# Maternal and child gluten intake and risk of type 1 diabetes: The Norwegian Mother and Child Cohort Study

**DOI:** 10.1101/19001883

**Authors:** Nicolai A Lund-Blix, German Tapia, Karl Mårild, Anne Lise Brantsaeter, Pål R Njølstad, Geir Joner, Torild Skrivarhaug, Ketil Størdal, Lars C Stene

**Author notes:** Correspondence: Nicolai A Lund-Blix, Department of Chronic Diseases and Ageing, Norwegian Institute of Public Health, P.O. Box 222 Skøyen, NO-0213 Oslo, Norway. These authors contributed equally to this work (Ketil Størdal, Lars C Stene).

## Abstract

**OBJECTIVE:** To examine the association between maternal and child gluten intake and risk of type 1 diabetes in children.

**DESIGN:** Pregnancy cohort

**SETTING:** Population-based, nation-wide study in Norway

**PARTICIPANTS:** 86,306 children in The Norwegian Mother and Child Cohort Study born from 1999 through 2009, followed to April 15, 2018.

**MAIN OUTCOME MEASURES:** Clinical type 1 diabetes, ascertained in a nation-wide childhood diabetes registry. Hazard ratios were estimated using Cox regression for the exposures maternal gluten intake up to week 22 of pregnancy and child’s gluten intake when the child was 18 months old.

**RESULTS:** During a mean follow-up of 12.3 years (range 0.7-16.0), 346 children (0.4%) developed type 1 diabetes (incidence rate 32.6 per 100,000 person-years). The average gluten intake was 13.6 grams/day for mothers during pregnancy, and 8.8 grams/day for the child at 18 months of age. Maternal gluten intake in mid-pregnancy was not associated with the development of type 1 diabetes in the child (adjusted hazard ratio 1.02 (95% confidence interval 0.73 to 1.43) per 10 grams/day increase in gluten intake). However, the child’s gluten intake at 18 months of age was associated with an increased risk of later developing type 1 diabetes (adjusted hazard ratio 1.46 (95% confidence interval 1.06 to 2.01) per 10 grams/day increase in gluten intake).

**CONCLUSIONS:** This study suggests that the child’s gluten intake at 18 months of age, and not the maternal intake during pregnancy, could increase the risk of type 1 diabetes in the child.

**WHAT IS ALREADY KNOWN ON THIS TOPIC:** A national prospective cohort study from Denmark found that a high maternal gluten intake during pregnancy could increase the risk of type 1 diabetes in the offspring (adjusted hazard ratio 1.31 (95% confidence interval 1.001 to 1.72) per 10 grams/day increase in gluten intake). No studies have investigated the relation between the amount of gluten intake by both the mother during pregnancy and the child in early life and risk of developing type 1 diabetes in childhood.

**WHAT THIS STUDY ADDS:** In this prospective population-based pregnancy cohort with 86,306 children of whom 346 developed type 1 diabetes we found that the child’s gluten intake at 18 months of age was associated with the risk of type 1 diabetes (adjusted hazard ratio 1.46 (95% confidence interval 1.06 to 2.01) per 10 grams/day increase in gluten intake). This study suggests that the child’s gluten intake at 18 months of age, and not the maternal intake during pregnancy, could increase the child’s risk of type 1 diabetes.

## INTRODUCTION

Type 1 diabetes is a common disease in childhood, and the Nordic countries have some of the highest incidence rates in the world.(1) Type 1 diabetes results from an immune-mediated destruction of pancreatic beta cells eventually leading to complete and lifelong dependence on exogenous insulin.(2) Although genetic susceptibility variants play a role for the development of type 1 diabetes, increased incidence rates over the past decades strongly suggest an important role of non-genetic factors.(1)

Gluten has been hypothesised to be one of the environmental factors involved in the development of type 1 diabetes.(3) It has been found in animal models and in vitro studies that gluten could have an effect on the immune system by increasing the proinflammatory cytokine production, or lead to dysbiosis of the gut microbiota.(3)

Most of the prospective studies in humans that have examined aspects of gluten intake as a risk factor for type 1 diabetes focus on age at introduction of cereals or gluten containing foods in infancy from high-risk cohorts with inconsistent results.(4-11) Two small studies reported no association between maternal intake of gluten containing foods in pregnancy and development of islet autoimmunity,(12, 13) but a recent large cohort study from Denmark found that a high maternal gluten intake during pregnancy could increase the risk of type 1 diabetes in the offspring.(14) No studies have investigated the relation between gluten intake by both the mother during pregnancy and the child in early life, and the risk of type 1 diabetes in the child.

Our objective was to examine the association between the maternal gluten intake during pregnancy, child’s gluten intake at age 18 months, and the risk of type 1 diabetes in the child.

## METHODS

### Participants and study design

We included participants in the Norwegian Mother and Child Cohort Study (MoBa).(15) MoBa is a prospective nation-wide population-based pregnancy cohort of pregnancies in Norway during 1999-2009. The women consented to participation in 41% of the pregnancies. A total of 86,306 children of whom 346 developed type 1 diabetes were included in the analysis (Figure 1).

**Figure 1.**
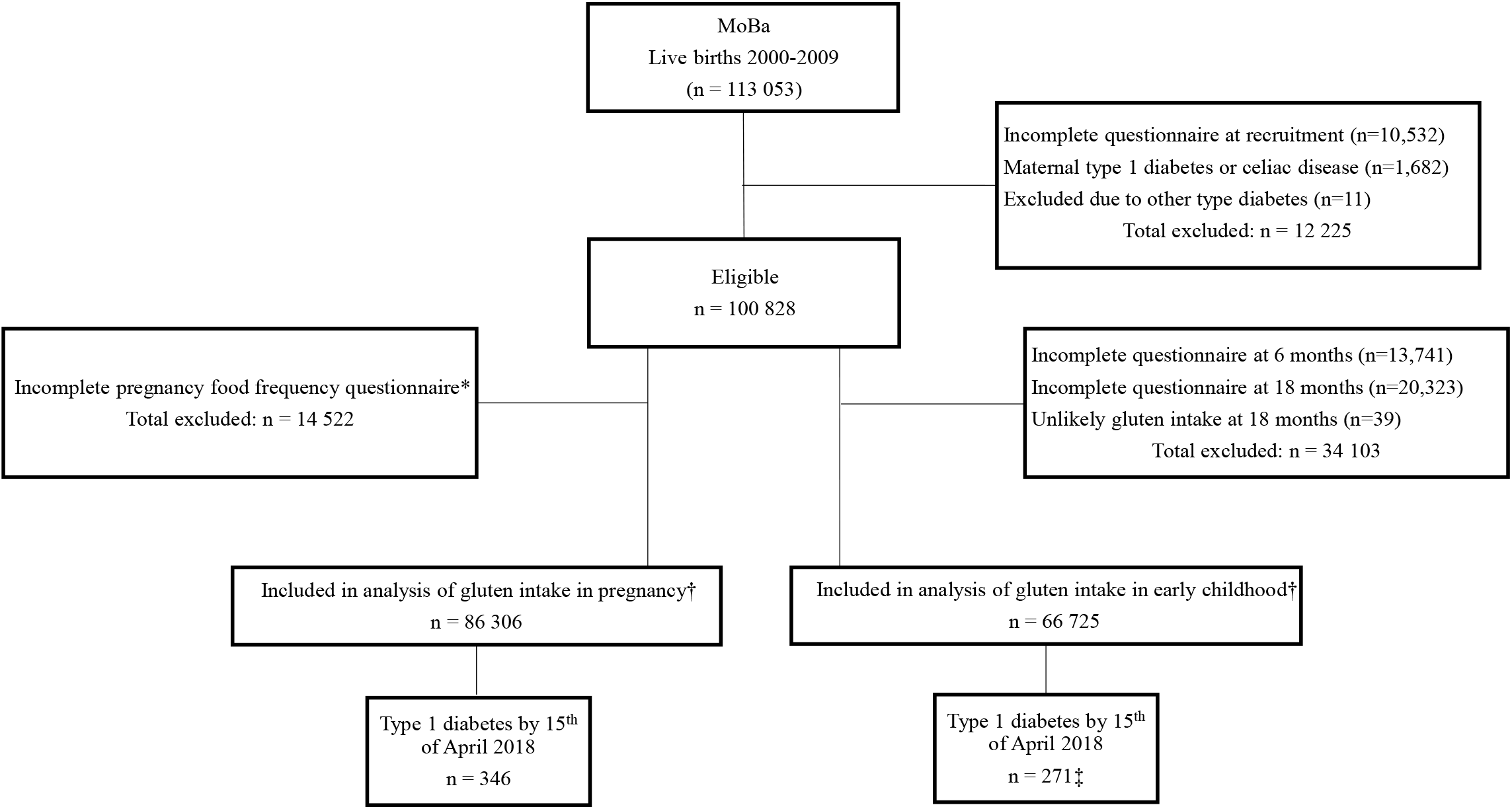
Flow-chart of the study cohort. * The food frequency questionnaire was introduced in 2002 (response rate 92%). † In the analysis including both pregnancy and childhood gluten intake, there was 62 900 participants with valid data (255 of these were type 1 diabetes cases). ‡ Nine cases where excluded from the analyses due to having the outcome type 1 diabetes prior to the exposure measurement at 18 months.

The current study is based on version VIII of the quality-assured data files. We used exposure information from questionnaires (available in English translation at https://www.fhi.no/en/studies/moba/for-forskere-artikler/questionnaires-from-moba) at week 22 of pregnancy and child age 18 months. Data was linked to the Medical Birth Registry of Norway and the Norwegian Patient Register using the personal identity number assigned to all Norwegian residents.

### Outcome: type 1 diabetes

We used time to clinical diagnosis of type 1 diabetes in the child as the outcome. Data on the child’s type 1 diabetes was obtained from the Norwegian Childhood Diabetes Registry (16) and the Norwegian Patient Registry. Nine cases of type 2- or monogenic diabetes were excluded from the study. A total of 346 children with type 1 diabetes with valid information on maternal gluten intake at week 22 of pregnancy and 271 with valid information on child’s gluten intake at 18 months of age were identified during follow-up (Figure 1). Participants excluded due to missing exposure information had an incidence of type 1 diabetes that did not differ from the incidence among those included in the current analysis (Supplementary Figure 1).

### Exposures: Gluten intake during pregnancy and in the child at age 18 months

We derived the amount (g/day) of gluten intake from a semi-quantitative food frequency questionnaire at week 22 of pregnancy and from a questionnaire completed by the guardian when the child was 18 months old (Supplementary Figure 2). For the food frequency questionnaire at week 22 of pregnancy we derived the average protein intake (g/day) from gluten containing flour or grains from the MoBa food database. The food frequency questionnaire covered the period up to week 22 of pregnancy, and has been validated for food and nutrient intake.(17) To estimate the child’s gluten intake we used a questionnaire completed at 18 months covering the frequency of wheat-, rye- and barley-containing food intake. Portion sizes were obtained from a published report (18) and product labels. We estimated the average protein intake (g/day) from gluten containing flour or grains by using recipes and nutritional contents of food items from the Norwegian Food Composition Table (19) in addition to traditional recipes and ingredient lists from product labels. The average gluten intake (grams/day) was estimated by using a conversion factor of 0.75 in accordance with most studies.(20-22) The conversion factor of 0.75 is based on conversion factors of 0.80 for wheat, 0.65 for rye, and 0.50 for barley,(22-24) and wheat being the most widely used grain in the different food products assessed.

We excluded from the analyses children with an unlikely high intake of gluten at 18 months of age (>35 g/day, n=39, Figure 1). We categorised maternal gluten intake during pregnancy and child’s intake at 18 months of age into percentiles (<10, 10-20, 20-50, 50-80, 80-90, ≥90) for comparison with the recently published study from Denmark.(14) Characteristics of participants are shown in Table 1 (Characteristics for participants with valid information on child’s gluten intake at 18 months of age is shown Supplementary Table 1). Characteristics of excluded participants due to missing exposure data are presented in Supplementary Table 2.

**Table 1.**
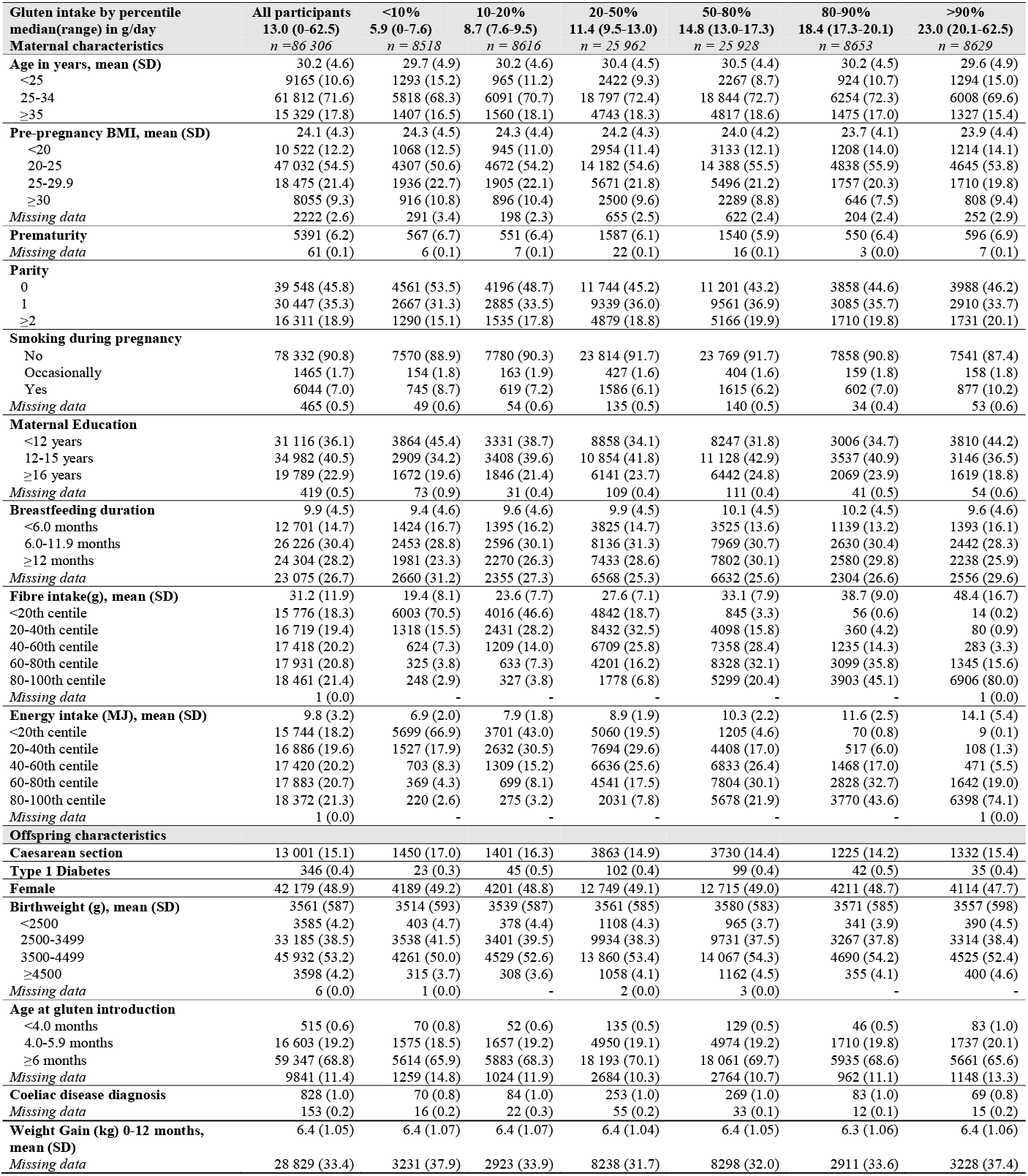
Characteristics of the study participants included in the analysis of maternal gluten intake during pregnancy and risk of type 1 diabetes in the child

**Table 2.** Association between maternal gluten intake during pregnancy (n = 86 306) or the child’s intake at 18 months (n = 66 725) and the risk of type 1 diabetes in the child

| Gluten intake | Cases | Incidence rate<br>(per 100 000) | Hazard ratio (95% CI) of type 1 diabetes |  |  |
| --- | --- | --- | --- | --- | --- |
|  |  |  | Unadjusted | Adjusted model 1* | Adjusted model 2†<br>(primary model) |
| Maternal/<br>Pregnancy |  |  |  |  |  |
| Continuous,<br>per 10 g/day<br>increase | 346 | 36.6 | 1.04 (0.86 - 1.27) | 1.02 (0.81 - 1.30) | 1.02 (0.73 - 1.43) |
| By category |  |  |  |  |  |
| <7.6 g/day | 23 | 22.3 | Ref. | Ref. | Ref. |
| 7.6-9.5 g/day | 45 | 42.6 | 1.90 (1.13 - 3.18) | 1.90 (1.13 - 3.20) | 1.71 (0.94 - 3.09) |
| 9.5-13.0 g/day | 102 | 32.0 | 1.42 (0.89 - 2.27) | 1.44 (0.90 - 2.31) | 1.37 (0.79 - 2.37) |
| 13.0-17.3 g/day | 99 | 31.1 | 1.39 (0.87 - 2.22) | 1.39 (0.84 - 2.29) | 1.23 (0.67 - 2.26) |
| 17.3-20.1 g/day | 42 | 39.5 | 1.76 (1.04 - 2.96) | 1.81 (1.04 - 3.16) | 1.50 (0.75 - 2.98) |
| >20.1 g/day | 55 | 32.7 | 1.45 (0.85 - 2.49) | 1.44 (0.78 - 2.63) | 1.60 (0.75 - 3.39) |
| P<sub>trend</sub> |  |  | 0.43 | 0.45 | 0.53 |
| Child/<br>18 Months of life |  |  |  |  |  |
| Continuous,<br>per 10 g/day<br>increase | 262 | 35.9 | 1.44 (1.05 - 1.98) | 1.44 (1.05 - 1.98) | 1.46 (1.06 - 2.01) |
| By category |  |  |  |  |  |
| <4.8 g/day | 19 | 26.3 | Ref. | Ref. | Ref. |
| 4.8-5.8 g/day | 25 | 34.3 | 1.32 (0.72 - 2.44) | 1.32 (0.72 - 2.43) | 1.30 (0.71 - 2.40) |
| 5.8-8.2 g/day | 77 | 34.9 | 1.35 (0.81 - 2.26) | 1.34 (0.80 - 2.25) | 1.33 (0.80 - 2.23) |
| 8.2-11.4 g/day | 76 | 34.7 | 1.25 (0.74 - 2.10) | 1.24 (0.74 - 2.10) | 1.24 (0.74 - 2.09) |
| 11.4-13.5 g/day | 30 | 41.2 | 1.61 (0.89 - 2.90) | 1.60 (0.89 - 2.89) | 1.61 (0.89 - 2.91) |
| >13.5 g/day | 35 | 48.2 | 1.84 (1.04 - 3.27) | 1.85 (1.04 - 3.28) | 1.86 (1.05 - 3.30) |
| P<sub>trend</sub> |  |  | 0.05 | 0.05 | 0.04 |

\* Model 1 (as in Antvorskov et al.(14)): Adjusted for maternal age, pre-pregnant maternal body mass index, parity, smoking during pregnancy, education, caesarean section, breastfeeding, sex and energy intake.
† Model 2 (primary model): As model 1 with additional adjustment for birthweight, age at gluten introduction, prematurity, fibre intake, weight gain 0-12 months and child's or mothers gluten intake (mutually adjusted exposures).

### Other variables

From the Medical Birth Registry of Norway we obtained maternal age, parity, mode of delivery, child sex, birth weight and gestational age categorised as shown in Table 1. The MoBa recruitment questionnaire completed at week 18 of pregnancy provided information regarding maternal education, smoking, pre-pregnant body mass index (BMI), maternal coeliac disease diagnosis and maternal type 1 diabetes diagnosis. From MoBa questionnaires completed at child age 6 and 18 months we obtained information on breastfeeding duration and child age at the time of gluten introduction.(25) MoBa questionnaires completed at child age 7 or 8 years and register linkage to the Norwegian Patient Register (to the end of 2016) provided information on the child’s coeliac disease diagnosis.(26) The Norwegian Patient Register also provided information on parents’ medical codes indicative of coeliac disease and type 1 diabetes. We excluded children of mothers with type 1 diabetes or coeliac disease due to the influence on the maternal gluten intake, and potentially the child’s gluten intake (n=1682; Figure 1).

### Statistical analysis

We used Cox regression analysis to estimate hazard ratios with 95% confidence intervals. Follow-up time was counted from birth when analysing maternal gluten intake, and from child’s age 18 months when analysing child’s gluten intake, to type 1 diabetes diagnosis or end of follow-up (April 15, 2018). We found no evidence for violation of the proportional hazards assumption by visually assessing log-minus log plots or formally testing Schoenfeld residuals. We used robust cluster variance estimation to account for potential correlation among siblings in the cohort. We predefined statistical significance as p-values 0.05 or 95% confidence intervals for the hazard ratio not including 1.00. The primary analysis was further predefined to be the test for linear trend (per 10 grams increase in gluten intake per day) with covariates defined in model 2 (see below), and to be based on a dataset where we imputed missing covariates using multiple imputation with chained equations.(27) To test non-linearity we analysed gluten as a categorical variable using the same cut-offs as the Danish national prospective cohort study (Table 1 and Supplementary Table 1).(14)

In addition to unadjusted analyses, we decided a priori to adjust for variables that may be associated with the gluten intake and type 1 diabetes. Model 1 adjusted for covariates similar to the Danish national prospective cohort study (14): pre-pregnancy maternal body mass index, age, parity, smoking status, education, breastfeeding duration, caesarean section, energy intake and child’s sex. Model 2, our primary model, additionally adjusted for age at gluten introduction, birthweight, prematurity, weight gain 0-12 months, maternal fibre intake during pregnancy, and mutually adjusted maternal and child’s gluten intake.

In sensitivity analyses to assess the impact of imputing missing covariates we repeated the main analyses in those with complete covariate data. We also assessed the association of maternal fibre intake and gluten intake from refined grains during pregnancy and risk of type 1 diabetes in the child. Also, we assessed the impact of further adjusting our analyses for the child’s coeliac disease. All analyses were done in Stata Release 15 (College Station, Tx, USA).

### Patient and public involvement

We conducted the study on previously collected data without any patient involvement in development of the research question or outcome measures, nor in development of design, recruitment and conduct of the study. Patients were not asked to advice on interpretation or writing up of results. Results from MoBa is disseminated to study participants through a yearly newsletter sent to all participating families, press releases, and the study’s and the Norwegian Institute of Public Health’s web sites.

## RESULTS

A total of 346 children (0.4%) developed type 1 diabetes after a mean of 12.3 years (range 0.7-16.0) of follow-up (Figure 1). The mean age at diagnosis was 7.5 years (range 0.7-15.0). We had information on islet autoantibodies (toward insulin, glutamic acid decarboxylase, and IA2) at diagnosis from 76% the children who developed type 1 diabetes, and 92% were positive for at least one islet autoantibody. The overall incidence rate of type 1 diabetes per 100,000 person-years was 32.6. Mean maternal gluten intake during pregnancy was 13.6 g/day (standard deviation 5.2, median 13.0 g/day) and the child’s mean gluten intake at age 18 months was 8.8 g/day (standard deviation 3.6, median 8.2 g/day).

Gluten intake was lower in mothers who had a lower fibre and energy intake. Gluten intake was also lower in children of mothers who were younger, less educated and breastfed their child for a shorter duration (Supplementary Table 1). Children of mothers with a low gluten intake during pregnancy tended to have a lower gluten intake at 18 months, but the correlation was weak (Supplementary Figure 3).

### Maternal gluten intake during pregnancy and risk of type 1 diabetes in the child

Maternal gluten intake during pregnancy was not associated with risk of type 1 diabetes in the child (Table 2). For each 10 g/day increase of gluten intake the adjusted hazard ratio was 1.02 (95% confidence interval 0.73 - 1.43). While there was a tendency towards a non-linear association, with higher risk of type 1 diabetes in those with the next lowest and those with the next highest category of maternal gluten intake, the global likelihood ratio test did not support a significant non-linear association (p = 0.11).

### Gluten intake by the child at age 18 months and subsequent risk of type 1 diabetes

The child’s gluten intake at 18 months was significantly associated with increased risk of type 1 diabetes with a dose-response relationship (Table 2). For each 10 g/day increase of gluten intake the adjusted hazard ratio was 1.46 (95% confidence interval 1.06 to 2.01).

While the child’s gluten intake remained significantly associated with risk of type 1 diabetes after adjustment for maternal gluten intake during pregnancy, the suggestive association with maternal gluten intake in category 2 and 5 were blunted after adjustment for child’s gluten intake (Table 2).

### Additional analyses

Analyses of those with complete covariate data showed similar results as in the main analyses (Supplementary Table 3). We found no association between maternal dietary fibre intake or gluten intake from refined grains during pregnancy and risk of type 1 diabetes in the child (Supplementary table 4 and 5). In analyses with further adjustment for child coeliac disease our results were essentially unchanged (Supplementary Table 6).

## DISCUSSION

In this first study with both maternal and child gluten intake as exposures, we found that the child’s gluten intake at 18 months of age was associated with the risk of type 1 diabetes while maternal gluten intake during pregnancy was not.

### Strengths and weaknesses

The main strengths of the study are the prospective design with recruitment in pregnancy, large sample size, and linkage to national registries with high level of ascertainment. Missing data on the child’s gluten intake at 18 months of age was mainly due to loss of follow-up, which occurs in all cohorts based on voluntary participation and questionnaires. The similar risk of type 1 diabetes in those with or without complete data suggest that bias due to missing data was not a serious problem. Furthermore, multiple imputation analysis have further contributed to minimising any influence of bias due to missing data.(27)

The recently published Danish national prospective cohort study has been criticised for not including other dietary components related to gluten intake.(28) We included data on maternal dietary fibre intake and gluten intake from refined grains during pregnancy. We also adjusted for age at gluten introduction and infant weight gain.(29) We accounted for coeliac disease, both by excluding children of mothers with coeliac disease from the analyses and adjusting for child coeliac disease.

Our study is observational so we cannot exclude the possibility that unmeasured confounders have influenced the results. On the other hand, there are no established environmental aetiological factors that are obvious confounders.

We assessed gluten intake prospectively, but imprecisions in the estimations are likely, and we only had information regarding gluten intake for the mother in the first half of pregnancy and the child at 18 months age. Nevertheless, our estimated gluten amount was similar to that from other cohorts using similar or more detailed dietary assessment methods, both for adults (14) and children.(30-33) Uncertainty in assessment of gluten intake from questionnaires is unlikely to depend on future type 1 diabetes, and any bias is likely to be non-differential when we have accounted for coeliac disease in the child and in the mother. The large majority of type 1 diabetes cases were diagnosed long after 18 months, so the potential for reverse causation in our study is likely to be small.

Self-selection among pregnant women in recruitment and loss to follow-up could result in a non-representative analysis sample. MoBa participants and those who continue to participate differ from the background population by higher education and maternal age and lower frequency of daily smoking.(34) The selection has been shown to play a minor role in association studies in MoBa.(34) Adjusting for maternal education, age and smoking during pregnancy did not change our results substantially, suggesting that selection bias is unlikely to have had a major influence on our findings.

### Comparison with previous studies

Our results on maternal gluten intake in pregnancy are not in line with those from the only previous study to assess this,(14) despite remarkable similarities in characteristics, maternal gluten intake and methodology. A possible explanation for the discrepancy could be that we were able to exclude participants based on maternal coeliac disease, while the other study did neither exclude nor adjust for this in their analyses. Two studies of high-risk children investigating maternal intake of gluten containing foods in pregnancy and development of islet autoimmunity reported no significant association.(12, 13) Of note, both the exposure (intake of gluten containing cereals, rather than amount of gluten) and outcome was different from our study, and the number of children with outcome was small in these high-risk cohorts. A recent analysis of data on estimated amount of gluten intake in children from the high-risk cohort DAISY found no significant association between the child’s gluten intake and progression from islet autoimmunity to type 1 diabetes.(35) Again, the apparent inconsistency with our result may have been due to studies investigating different outcomes (clinical type 1 diabetes vs progression) in a different population, or lack of power. Our results are probably generalizable to other industrialised countries, but may not be applicable to populations of non-European origin.

## Conclusions and policy implications

This study suggests that the child’s gluten intake at 18 months of age, and not the maternal intake during pregnancy, could increase the risk of type 1 diabetes. The results from a similar Danish national prospective cohort study reporting that a high maternal gluten intake during pregnancy could increase the risk of type 1 diabetes in the child (14) was not replicated in our cohort, suggesting that there is no causality. This could also be the case for our finding of the child’s gluten intake at 18 months as a possible risk factor for type 1 diabetes later in life, which needs to be replicated in other large cohorts or intervention studies.

The child’s gluten intake at 18 months of age could possibly increase the risk of type 1 diabetes through several mechanisms related to the immune system.(3) It has been shown that increased gut permeability which facilitates for abnormal absorption of macromolecules is associated with type 1 diabetes, and detectable before clinical onset.(36) In a recently published study from The Environmental Determinants of Diabetes in the Young (TEDDY) cohort small changes in abundance of bacterial genera was found in cases with type 1 diabetes compared to controls, where controls had a higher abundance of bacterial genera that might be indicative of enhanced gut integrity.(37) The child’s diet in early life is more important for microbiota development in the child than maternal diet during pregnancy, and our assessment of gluten intake at 18 months of age is in the transitional phase of the developing microbiome before the stable phase observed after 30 months of age.(37) The median age at seroconversion to islet autoimmunity preceding type 1 diabetes is around 24 months of age.(38)

Our observations may motivate future interventional studies with reduced gluten intake to establish whether there is a true causal association between amount of gluten intake in the child’s early diet and type 1 diabetes in susceptible individuals. There is no current evidence for mothers to reduce gluten intake during pregnancy. However, ours and other currently available data should be interpreted with caution, given the few prospective studies in this field with conflicting results and a lack of randomised intervention studies.

In conclusion, we found that a higher intake of gluten in the child’s diet at 18 months, but not in the maternal diet during pregnancy, was associated with an increased risk of type 1 diabetes in the child.

## Data Availability

Codes for the statistical analyses are available upon request. Access to data can be obtained by sending an application to The Norwegian Mother and Child Cohort Study (, information website: https://www.fhi.no/en/op/data-access-from-health-registries-health-studies-and-biobanks/data-from-moba/moba-research-data-files/)

## ACKNOWLEDGEMENTS

We are grateful to all the participating families in Norway who take part in this on-going cohort study.

## Contributors

LCS and KS conceptualised and designed the study, acquired pregnancy cohort data, and obtained funding. NAL, LCS and KS performed the literature search. GT, NAL, KS and LCS analysed the data and drafted the initial manuscript. ALB acquired dietary data from the Norwegian Mother and Child Cohort Study dietary database. NAL quantified gluten intake. TS, GJ and PRN acquired incident type 1 diabetes data. All authors interpreted the results, reviewed and revised the manuscript. LCS and KS are the guarantors and accepts full responsibility for the work and the conduct of the study, had access to the data, and controlled the decision to publish. The corresponding author attests that all listed authors meet authorship criteria and that no others meeting the criteria have been omitted.

## Copyright/license for publication

The Corresponding Author has the right to grant on behalf of all authors and does grant on behalf of all authors, a worldwide licence to the Publishers and its licensees in perpetuity, in all forms, formats and media (whether known now or created in the future), to i) publish, reproduce, distribute, display and store the Contribution, ii) translate the Contribution into other languages, create adaptations, reprints, include within collections and create summaries, extracts and/or, abstracts of the Contribution, iii) create any other derivative work(s) based on the Contribution, iv) to exploit all subsidiary rights in the Contribution, v) the inclusion of electronic links from the Contribution to third party material where-ever it may be located; and, vi) licence any third party to do any or all of the above.

## Ethical approval

The establishment and data collection in MoBa was previously based on a licence from the Norwegian Data Inspectorate and approval from The Regional Committee for Medical Research Ethics, and it is now based on regulations related to the Norwegian Health Registry Act. All participants provided written informed consent. The present study was approved by The Regional Committee for Medical Research Ethics.

## Transparency declaration

The lead author affirms that the manuscript is an honest, accurate, and transparent account of the study being reported; that no important aspects of the study have been omitted; and that any discrepancies from the study as originally planned have been explained.

